# When advantage turns into risk: disentangling landscape and behavioural drivers of socioeconomic inequality in Lyme disease risk, Glasgow as a case study

**DOI:** 10.64898/2026.05.18.26353476

**Authors:** Sara L. Gandy, Grace Plahe, Jessica Hall, Kirsty Watkinson, Swapna Guntupalli, David Johnson, Richard Birtles, Sally Mavin, Lucy Gilbert

## Abstract

**Introduction:** Socioeconomic deprivation is often associated with poorer health outcomes, but some studies suggest the opposite for Lyme disease. Here we test two hypotheses to explain this: differences in (i) local landcover of high risk habitats such as woodlands (landscape hypothesis) and (ii) outdoor recreation in such habitats (behaviour hypothesis).

**Methods:** We analysed reported Lyme disease incidence data for 824 data zones in the city of Glasgow, UK, against deprivation rank (based on indicators relating to income, employment, health, education, crime and housing). We then tested how these relate to woodland cover and indices of urban greenspace usage (per capita and per ha of greenspace). Additionally, we measured Lyme disease hazard (density of infected ticks) in 32 greenspaces and tested relationships with deprivation, woodland and greenspace usage.

**Results:** More advantaged data zones (data zones with low deprivation rank) had higher Lyme disease incidence. These areas had more woodland and woodland cover was positively correlated with both Lyme disease incidence and hazard. Deprivation did not correlate with greenspace usage, nor did greenspace usage correlate with Lyme disease incidence. Intensely used greenspaces had lower infected tick densities, consistent with a human disturbance effect on wildlife that carry ticks.

**Conclusions:** Differences in woodland cover, but not outdoor recreation behaviour, can help explain our finding of higher Lyme disease incidence in more advantaged areas. However, to further test the behaviour hypothesis, we need more detailed data on outdoor recreation activity per capita both locally and in rural areas, as well data on mitigation behaviours.

## Introduction

Public health outcomes are often linked to socioeconomics, with disadvantaged populations suffering systematic inequalities amplifying burdens of diseases such as autoimmune and cardiovascular diseases, and some mosquito-borne diseases.^1–4^ In contrast, recent evidence suggests that higher socioeconomic groups experience higher incidence of tick-borne diseases such as Lyme disease in both North America^5–7^ and Europe.^8^

Ecological transmission cycles of tick-borne diseases are complex, involving interactions between ticks, pathogens and their respective vertebrate hosts, as well as environmental factors.^9^ In Europe, *Ixodes ricinus* is the primary vector for *Borrelia burgdorferi* sensu lato (s.l.), the bacterial complex causing Lyme disease.^10^ The three active life stages of *I. ricinus* each take a blood meal from a vertebrate host in order to develop to the next life stage (larvae and nymphs) or to lay eggs (adult females).^9^

Reported Lyme disease incidence depends on a combination of the environmental hazard (the density of infected *I. ricinus* nymphs active in the environment), people’s exposure to infected ticks, their post-exposure or mitigation behaviour, medical diagnosis and reporting^11^ (online supplementary figure 1). Socioeconomic inequalities in Lyme disease incidence must therefore relate to differences in one or more of these factors. For example, areas of higher socioeconomic status might have higher local environmental Lyme disease hazard, due to more high risk habitats such as woodlands.^5,7^ Woodlands harbour high densities of *I. ricinus* tick hosts, such as deer, as well as high densities of *B. burgdorferi* s.l. transmission hosts, such as rodents and birds. In addition, they also provide a mild, humid micro-climate aiding tick activity and survival.^9,12,13^ Therefore, woodlands are the habitat most commonly associated with environmental Lyme disease hazard.^14,15^

Furthermore, socioeconomic status can be associated with certain behaviours that could affect people’s exposure: higher socioeconomic groups in Europe participate in more leisure-time physical activities^16^ and people living in more deprived urban areas are less likely to regularly use greenspaces.^17^ This is due to fewer high quality greenspaces available^18^ in addition to safety concerns and accessibility issues related to available greenspaces^19^ There is little evidence that mitigation behaviours (using tick repellents, tick checks) correlate with education level or income socioeconomic variables,^20^ and these were thus not considered in this study.

In this study, we test hypotheses relating to the environment and to pre-exposure behaviour in a large UK metropolitan area and its surrounding peri-urban and rural fringe (figure 2). We predict that Lyme disease incidence will be higher in areas of higher socioeconomic status (prediction 1). We then test two hypotheses to explain this effect (figure 1):

**Figure 1.**
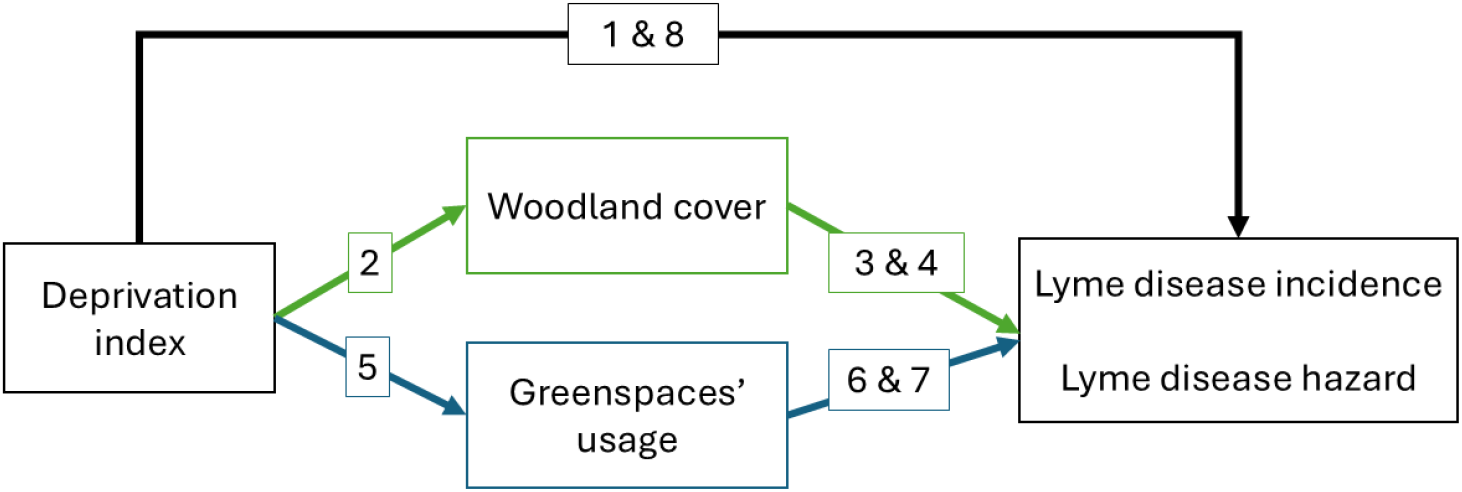
Conceptual diagram representing the hypothesised pathways tested in this study, linking socioeconomic status and Lyme disease incidence and environmental hazard. Links in green represent the Landscape Hypothesis and links in blue represent the Behaviour Hypothesis. Numbers represent each prediction tested.

**Figure 2.**
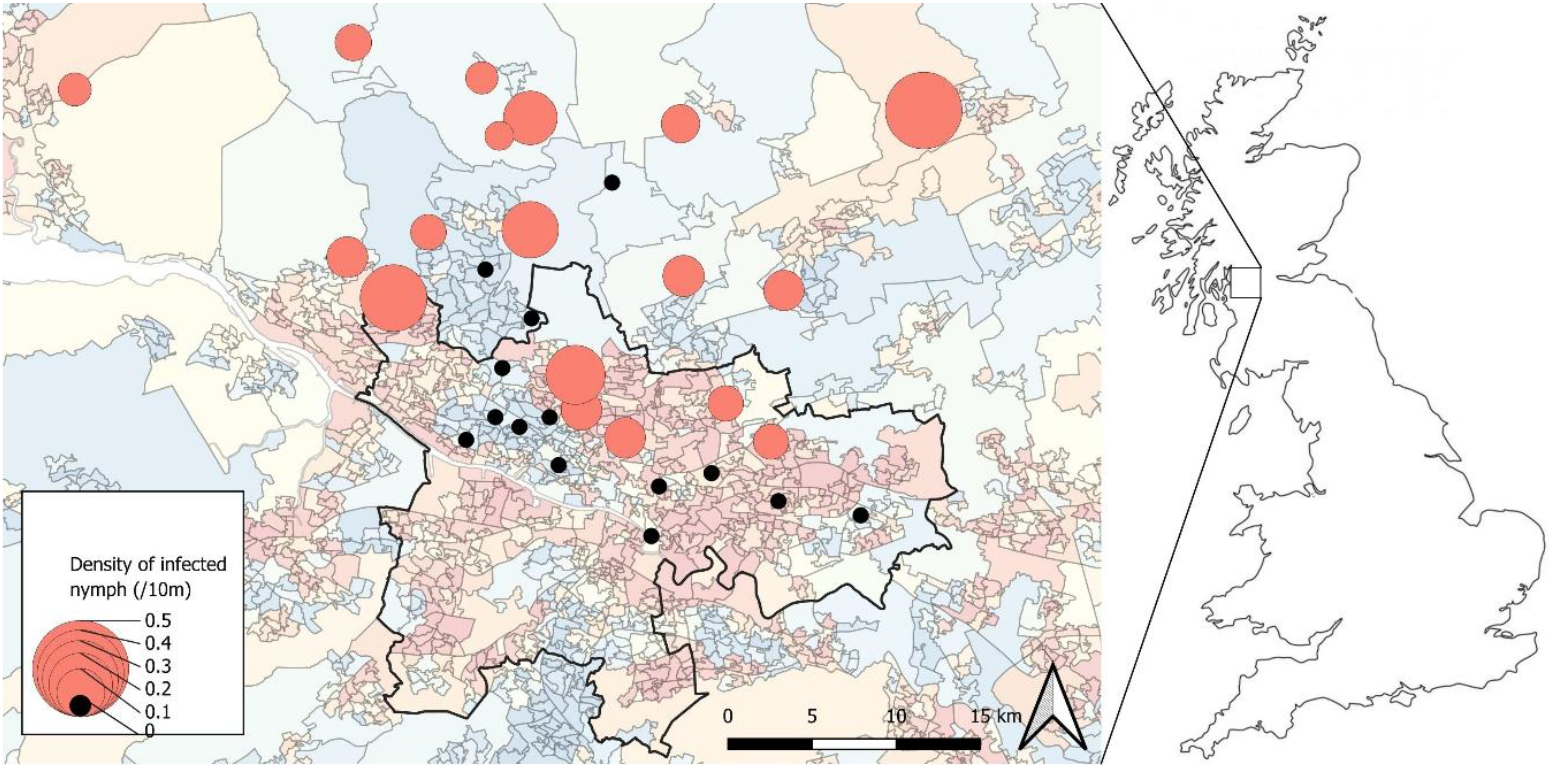
Map of the Greater Glasgow area showing the average deprivation rank from most deprived (dark red) to most advantaged (dark blue) for each data zone (polygons). Pink circles represent the 32 greenspaces at which we collected data on the environmental hazard of Lyme disease. The size of the circles represents the density of infected nymphs while black circles represent sites with no infected nymphs. The black line shows the Glasgow City Council boundary, within which we analysed Lyme disease incidence at the data zone level.

*The Landscape Hypothesis* is that people in urban areas of higher socioeconomic status (thereafter termed as “more advantaged areas”) are more likely to acquire infection because their local areas have more high risk habitats (i.e. woodlands). We therefore expect more advantaged areas to have higher woodland cover^21,22^ (prediction 2); and that woodland cover is positively associated with Lyme disease hazard (densities of infected nymphs) (prediction 3)^14,15^ and therefore, Lyme disease incidence (prediction 4).

*The Behaviour Hypothesis* is that people in more advantaged urban areas are more likely to become infected because they spend more time in high-risk habitats. We therefore predict higher usage of greenspaces in more advantaged areas^17–19^ (prediction 5); and higher greenspace usage should be associated with higher incidence of Lyme disease (prediction 6). As this hypothesis is based on differences in human behaviour rather than in environmental hazard, it is important to test whether greenspace usage also affects Lyme disease hazard (prediction 7).

Irrespective of woodland cover or greenspace usage, there could also be an association between socioeconomic status and environmental Lyme disease hazard (prediction 8), if there are consistent differences between socioeconomic status in other environmental characteristics that could affect tick and pathogen hosts, such as park management practices (e.g. grass cutting, leaf clearing).

## Materials and Methods

### Study design

The study was conducted in and around Glasgow, Scotland’s largest city, with an estimated population of 623,000 within the city council boundary and over 1 million in the contiguous urban area. The city encompasses over 3,500 hectares of greenspace and is characterised by pronounced socioeconomic inequalities (figure 2).^23,24^ For administrative purposes, the city is subdivided into 824 geographic units known as data zones, each having a unique deprivation rank, estimated by the Scottish Government based on seven indicators related to income, employment, health, education, geographic access to services, crime and housing (online supplementary table 1).^25^

We used two different approaches at different spatial scales: a Glasgow city-wide analysis using data for the 824 data zones to test our hypotheses linking socioeconomic status with Lyme disease incidence; and a finer scale analysis conducted at 32 greenspaces located inside, or within 14 km of, the city boundary (24 urban/suburban greenspaces, 8 rural woodlands). These sites were specifically chosen across both urban-rural and socioeconomic gradients to test our hypotheses linking socioeconomic status with landcover and Lyme disease environmental hazard (figure 2).

### Deprivation rank

#### Data Zone analysis

The deprivation rank for each data zone was obtained from the Scottish Index of Multiple Deprivation^25^ by averaging the ranks for income, employment, health, education, crime and housing (mean: 2165; median: 1708; range: 99-6236; figure 3). Lower values indicate greater deprivation and higher values indicate higher socioeconomic areas.

**Figure 3.**
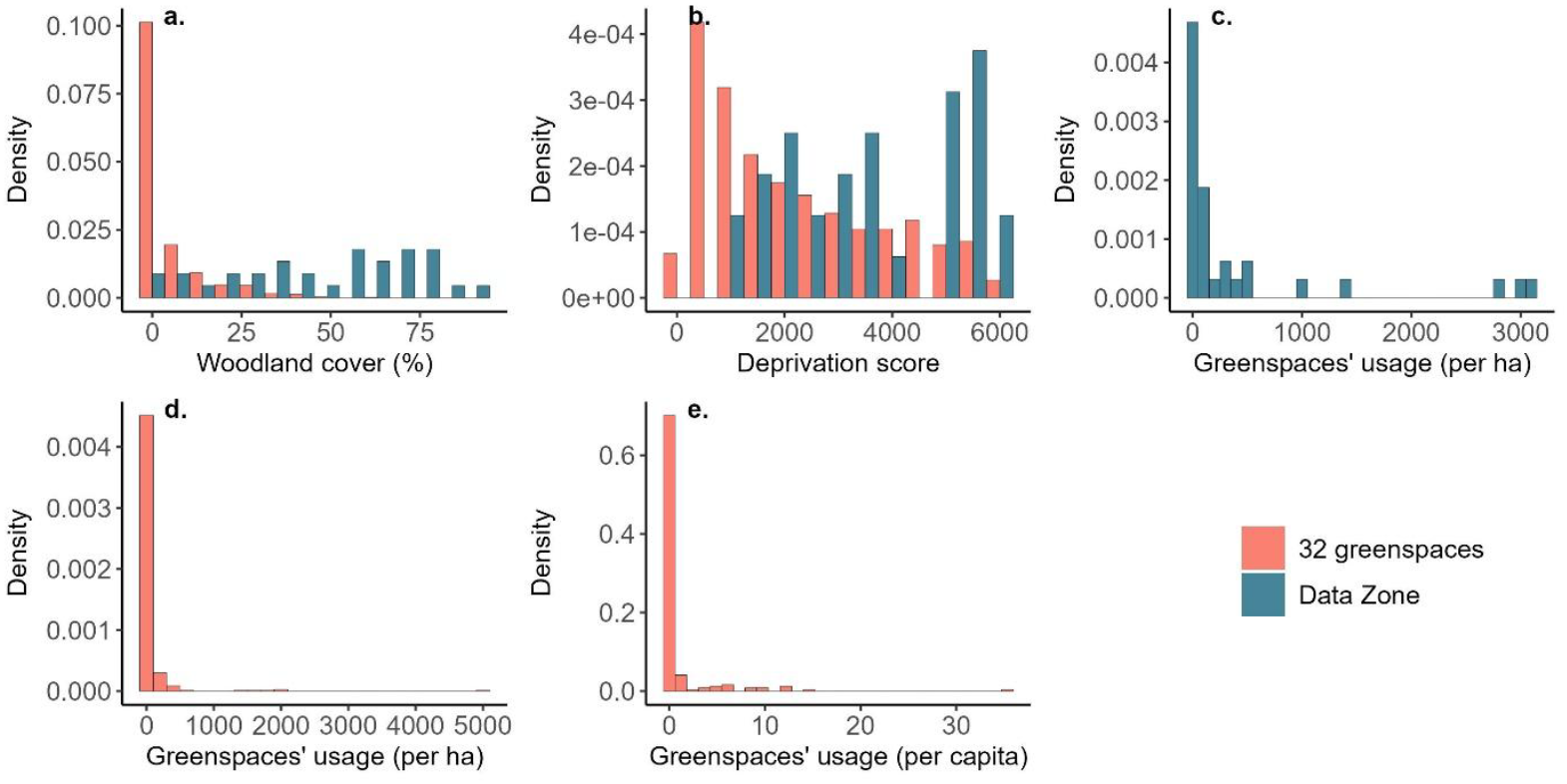
Frequency distribution of the different metrics: (a) proportion of woodland cover within each data zone (blue) and within each of the 32 greenspaces surveyed (orange); (b) average deprivation rank within each data zone (blue) and within a 1 km buffer from the edge of each of the 32 greenspaces surveyed (orange); (c) greenspace usage per ha of greenspace in the 32 greenspaces surveyed; (d) greenspace usage (cycling trips per ha of greenspace) at the data zone level (212 data zones with greenspaces >1.5 ha included); (e) greenspace usage (cycling trips per capita) at the data zone level (212 data zones with greenspaces >1.5 ha included).

#### 32 greenspaces analysis

Using the same deprivation dataset, we calculated the average deprivation rank within a 1 km wide buffer around the edge of each greenspaces (mean: 3631; median: 3556; range: 1032-6027; figure 3).

### Lyme disease case data (data zone analysis only)

We used laboratory confirmed Lyme disease cases, which consisted of patients that tested *B. burgdorferi* s.l. IgG and/or IgM positive via the internationally recognised standard two-tiered protocol.^26^ This represented 151 cases between 2018 and 2024 within the Glasgow City Council boundary. For analysis we used the number of cases per year for each of the 824 data zones (using R version 4.3.3, RCore Team), weighted by data zone population size.

### Environmental Lyme disease hazard (greenspaces analysis only)

Environmental Lyme disease hazard is the density of nymphal ticks infected with *B. burgdorferi* s.l., so we conducted surveys at the 32 greenspaces to estimate nymph density and analysed the nymphs for the presence of *B. burgdorferi* s.l.

#### Tick density

We estimated questing *I. ricinus* nymph density by surveying each of the 32 greenspaces twice between 2022 and 2024. We used a standard dragging method whereby a 1 m x 1 m woollen blanket was dragged over ground vegetation along 10 m transects and nymphal ticks were counted and collected.^27^ At each site visit, 15 transects were conducted. If fewer than 50 nymphs were collected, dragging was continued for a further hour to increase the sample size of ticks for pathogen screening. Tick surveys were conducted on dry days and at temperatures above 10 °C, to minimise the effects of rain and low temperatures on tick activity.^27^ Other variables that affect tick counts were recorded: air temperature, tree canopy closure^28^ and a ground vegetation height-density index,^27^ termed hereafter as “ground vegetation density”. We were not interested in these variables *per se*, but their effects needed to be taken into account in statistical models.

#### Borrelia burgdorferi s.l. identification

Each individual nymph collected was tested for the presence of *B. burgdorferi* s.l. by extracting DNA followed by PCR, using the method previously described^29^ (online supplementary text 1). For statistical analysis, Lyme disease hazard at each greenspace was modelled as the density of ticks positive for *B. burgdorferi* s.l., using the area surveyed as an offset.

### Woodland cover

#### Data zone analysis

We used landcover maps^30^ to calculate the proportion of woodland cover (deciduous and coniferous combined) for each data zone (mean: 4.7 %; median: 0.4%; range: 0-63.7 %; figure 3).

#### 32 greenspaces analysis

To test the link between socioeconomic status and woodland cover (prediction 2), we calculated the proportion of woodland cover within a 1 km buffer around the edge of each site, using landcover maps^30^ (mean: 14.5 %; median: 11.4%; range: 0.3-43.9 %; figure 3). To test the link between woodland cover and Lyme disease hazard (prediction 3), we used the proportion of woodland cover within each greenspace.

### Greenspace usage

#### Data zone analysis

We used all publicly accessible greenspaces greater than 1.5 ha within the Glasgow city council boundary: 282 greenspaces across 212 data zones. To represent relative usage of these 282 greenspaces, we used Strava cycling data for 2019.^31^ We used cycling data because walking data were not available; we assumed that relative cycling usage between the sites was representative of other types of usage. For analysis, we used the number of non-commuting cycling trips per urban greenspaces in 2019, to best reflect leisure activities (mean: 67.7; median: 0; range: 0-5000; figure 3). We modelled greenspace usage as (i) per unit area (cycling trips per ha of greenspace) as a proxy of how busy each greenspace is; and (ii) per capita (cycling trips in all greenspaces in a data zone divided per population) as a proxy of how much each person uses local greenspace.

#### 32 greenspaces analysis

We combined the Strava cycling data for 2019^31^ with two social media datasets (Flickr and All Trails) to create a usage index using a weighted data synthesis approach in Python.^32^ This approach weights each dataset using the principal of central tendency to reduce the impact of any extreme counts within each individual dataset and produces estimates of the total number of visits to each site divided by the area of the greenspace (mean: 454; median: 73; range: 0-3085; figure 3).

### Statistical analysis

Statistical analyses were performed in R (R Core Team version 4.3.3). Any potential spatial autocorrelation in the models’ residuals was accounted for by including Moran eigenvectors (calculated with a weights matrix based on queen contiguity and Monte Carlo randomization using 999 simulations) as independent variables when needed.^5^ This filtered out any spatial autocorrelation detected between the neighbourhood levels observations from the model residuals.^5^

#### Prediction 1-Association between socioeconomic status and Lyme disease incidence

This analysis was conducted at the data zone level. We used a negative binomial Generalised Linear Mixed Effect Model (GLMM) with the number of Lyme disease cases per year and per data zone as the response variable. The covariates were the average deprivation rank for each data zone, year and the Moran eigenvectors to account for spatial autocorrelation. An offset for the total population at the data zone level was included, and data zone was added as a random effect.

#### Prediction 2- Association between socioeconomic status and woodland cover

For the analysis at the data zone level, we used a negative binomial Generalised Linear Model (GLM) with the average deprivation rank for each data zone rounded to the nearest integer as the response variable. Covariates included the proportion of woodland cover for each data zone in its quadrative form and Moran eigenvectors to account for spatial autocorrelation. We also conducted this analysis at our 32 greenspaces; We used a negative binomial GLM with the average deprivation rank within a 1 km buffer around the edge of each greenspace, rounded to the nearest integer, as the response variable. The covariate was the proportion of woodland cover within the same 1 km buffer around the edge of each site.

#### Prediction 3- Association between Lyme disease hazard and woodland cover

This analysis was conducted across our 32 greenspaces. We used data at the site-visit level (2 visits per greenspace), with our response variable being the number of *I. ricinus* nymphs infected with *B. burgdorferi* s.l., using with an offset for the area surveyed (e.g. 5 infected nymphs for 150 m^2^ surveyed). We used a negative binomial GLMM and the covariates included were Julian day, temperature, average vegetation density, canopy closure, park’s size in its quadratic form and the proportion of woodland cover within each site. Site was added as a random effect and we performed model selection using the dredge function from the MuMIn package,^33^ selecting the model with the lowest AICc. The selected model only included woodland cover within each site and temperature.

#### Prediction 4-Association between Lyme disease incidence and woodland cover

We conducted this analysis at the data zone level, using a negative binomial GLM with the number of Lyme disease cases per data zone as the response variable and an offset for the total population in each data zone. Covariates included the proportion of woodland cover for each data zone in its quadrative form and Moran eigenvectors to account for spatial autocorrelation.

#### Prediction 5-Association between socioeconomic status and greenspace usage

For the analysis at the data zone level, we ran two negative binomial GLMs. The first one used the number of non-commuting cycling trips per ha of greenspaces for each of the 212 data zones that had greenspaces over 1.5 ha, rounded to the nearest integer, as the response variable. The second model used the total number of non-commuting cycling trips divided by the population size (i.e. per capita) of the data zone for each of the 212 data zones as the response variable. For both models, the covariates were the deprivation rank for each data zone and Moran eigenvectors to account for spatial autocorrelation. For the analysis across our 32 greenspaces, we used a negative binomial GLM, with the greenspace usage index generated (using Strava, Flickr and All Trails) rounded to the nearest integer as the response variable. The average deprivation rank within a 1 km buffer from the edge of each greenspace was our covariate.

#### Prediction 6-Association between Lyme disease incidence and greenspace usage

We conducted the analysis at the data zone level. Due to convergence issues when trying to use the number of Lyme disease cases per data zone as the response variable, we had to transform it into a binary variable; if a data zone had at least one Lyme disease case, it scored a “1”, while data zones with no cases scored a “0”. When then used two binomial GLMs with a logit link, using either the number of non-commuting cycling trips per ha of greenspace in each data zone as the covariate (model 1) and the number of non-commuting cycling trips per capita in each data zone as the covariate (model 2), including the total population as an offset in both models.

#### Prediction 7-Association between Lyme disease hazard and greenspace usage

We used data from our 32 greenspaces at the site-visit level (2 visits per site) and our response variable was the number of *I. ricinus* nymphs infected with *B. burgdorferi* s.l. with an offset for the area surveyed (e.g. 5 infected nymphs for 150 m^2^ surveyed). We used a negative binomial GLMM with the covariates included being Julian day, temperature, average vegetation density, canopy closure, park’s size in its quadratic form and our greenspace usage index. Site was added as a random effect and we performed model selection using the dredge function from the MuMIn package,^33^ selecting the model with the lowest AICc. The selected model only included our greenspace usage index and temperature.

#### Prediction 8-Association between socioeconomic status and Lyme disease hazard

we used data at the site-visit level (2 visits per site) from our 32 greenspaces and our response variable was the number of *I. ricinus* nymphs infected with *B. burgdorferi* s.l. with an offset for the area surveyed (e.g. 5 infected nymphs for 150 m^2^ surveyed). We used a negative binomial GLMM with the covariates included being Julian day, temperature, average vegetation density, canopy closure, park’s size in its quadratic form and the average deprivation rank within a 1 km buffer around the edge of each site. Site was added as a random effect and we performed model selection using the dredge function from the MuMIn package,^33^ selecting the model with the lowest AICc. The selected model only included our temperature however, we added deprivation rank to generate delta AICc and predictions.

## Results

Statistical outputs for the focal parameters for each prediction are detailed below. Outputs for the additional factors that needed to be taken into account but were not of specific interest (temperature, canopy cover, ground vegetation density, Julian day, park size) are available in online supplementary table 2.

Prediction 1: More advantaged data zones had significantly higher incidence of Lyme disease than did more deprived data zones (IRR = 1.24, 95% CI: 1.03–1.49, p=0.02, figure 4a, table 1).

**Table 1.** Outputs from the GLMs and GLMMs described in Table 1 testing each numbered prediction. Outputs are shown for the key variables of interest: deprivation index, woodland cover, greenspace usage. The full model output summaries are available in online supplementary table 2. Delta AICc indicates the change in AICc when the variable is removed from the model (positive is worse fit; negative is better fit).

| Level | Response | Covariate | Estimate | Std. error | z-value | p-value | Delta AICc |
| --- | --- | --- | --- | --- | --- | --- | --- |
| **1- Association between socioeconomic status and Lyme disease incidence** |  |  |  |  |  |  |  |
| Data Zone | Number of cases | Deprivation rank | 0.21 | 0.09 | 2.25 | 0.02 | 3.0 |
| **2- Association between socioeconomic status and woodland cover** |  |  |  |  |  |  |  |
| Data Zone | Deprivation rank | Woodland cover | -0.01 | 0.01 | -2.05 | 0.04 | 2.5 |
|  |  | Woodland cover<sup>2</sup> | 0.0005 | 0.001 | 3.43 | <0.001 | 1.0 |
| 32 greenspaces | Deprivation rank 1 km buffer | Woodland cover 1 km buffer | 0.01 | 0.01 | 2.13 | 0.03 | 2.0 |
| **3- Association between Lyme disease hazard and woodland cover** |  |  |  |  |  |  |  |
| 32 greenspaces | Lyme disease hazard | Woodland cover within each site | 0.78 | 0.37 | 2.09 | 0.04 | 1.8 |
| **4-Association between Lyme disease incidence and woodland cover** |  |  |  |  |  |  |  |
| Data Zone | Number of cases | Woodland cover | -0.47 | 0.19 | -2.48 | 0.01 | 3.8 |
|  |  | Woodland cover<sup>2</sup> | 0.44 | 0.15 | 3.00 | 0.003 | 5.8 |
| **5-Association between socioeconomic status and greenspace usage** |  |  |  |  |  |  |  |
| Data Zone | Cycling trips per ha of greenspace | Deprivation rank | -0.24 | 0.47 | -0.51 | 0.61 | -1.8 |
| Data Zone | Cycling trips per capita | Deprivation rank | -0.21 | 0.57 | -0.38 | 0.71 | -2.02 |
| 32 greenspaces | Greenspace usage | Deprivation rank 1 km buffer | -0.50 | 0.37 | -1.34 | 0.2 | -0.7 |
| **6-Association between Lyme disease incidence and greenspace usage** |  |  |  |  |  |  |  |
| Data Zone | Number of cases | Cycling trips per ha of greenspaces | -0.31 | 0.34 | 0.92 | 0.36 | -7.2 |
| Data Zone | Number of cases | Cycling trips per capita | -0.17 | 0.29 | -0.58 | 0.56 | -1.6 |
| **7-Association between Lyme disease hazard and greenspace usage** |  |  |  |  |  |  |  |
| 32 greenspaces | Lyme disease hazard | Greenspace usage | -1.48 | 0.57 | -2.60 | 0.009 | 5.1 |
| **8-Association between socioeconomic status and Lyme disease hazard** |  |  |  |  |  |  |  |
| 32 greenspaces | Lyme disease hazard | Deprivation rank 1 km buffer | -0.29 | 0.29 | -0.98 | 0.33 | -1.4 |

**Figure 4.**
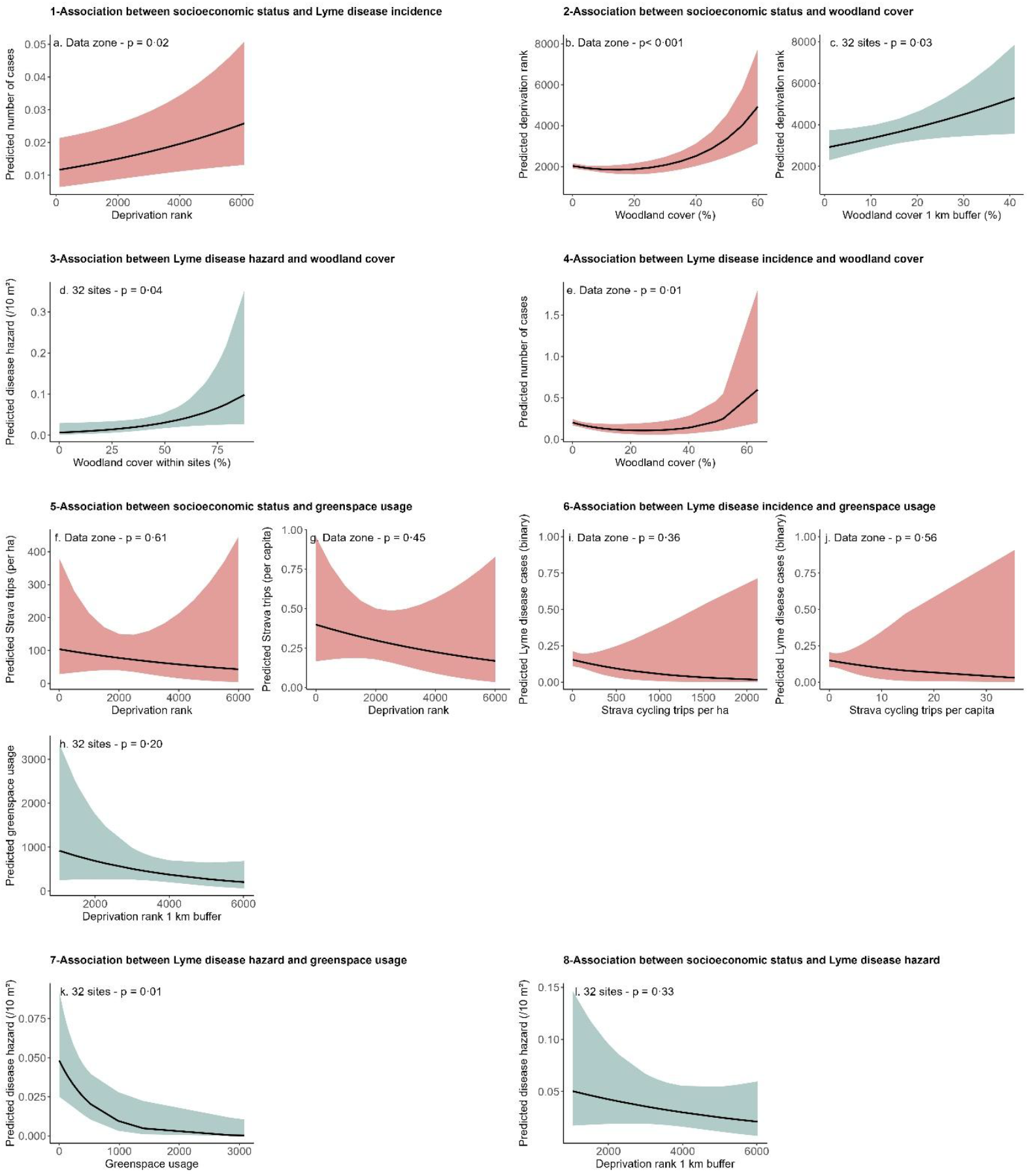
Visual outputs from GLMs and GLMMs testing the predictions for the associations between: 1 - socioeconomics and Lyme disease incidence (a); 2 - socioeconomics and woodland cover in data zones (b) and around 32 greenspaces (c); 3 and 4 - woodland cover and Lyme disease hazard (d) and Lyme disease incidence (e); 5 - socioeconomics and greenspace usage (Strava cycling trips) per ha (f) and per capita (g) at the data zone level, and greenspace usage index in 32 greenspaces (h); 6 – Lyme disease incidence and greenspace usage per ha (i) and per capita (j); 7 – greenspace usage index in 32 greenspaces and Lyme disease hazard (k); 8 – socioeconomics and Lyme disease hazard (l). Red represents analyses of the 824 data zones and blue of the 32 greenspaces. Shaded areas represent 95% confidence intervals.

### Landscape Hypothesis

Prediction 2: Deprivation rank was positively associated with woodland cover at both the data zone level (p<0.001, Figure 3b, Table 1) and in a 1 km buffer around the 32 greenspaces (IRR = 1.01, 95% CI: 1.00–1.03, p = 0.03, Figure 3c, Table 1).

Predictions 3 and 4: woodland cover was positively associated with the environmental hazard of Lyme disease within the 32 greenspaces (IRR = 2.17, 95% CI: 1.05–4.50, p = 0.04; Figure 3d, Table 1), and with Lyme disease incidence at the data zone level (p = 0.01, Figure 3e, Table 1).

### Behaviour Hypothesis

Prediction 5: Deprivation rank was not correlated with greenspace usage (cycling trips recorded on Strava) at the data zone level, neither per ha of greenspace (IRR = 0.79, 95% CI: 0.31-1.97, p = 0.61, Figure 3f, Table 1) or per capita (IRR = 0.79, 95% CI: 0.43-1.45, p = 0.45, Figure 3g, Table 1); nor with greenspace usage index in the 32 greenspaces (IRR = 0.61, 95% CI: 0.29-1.26, p = 0.2, Figure 3h, Table 1).

Prediction 6: There was no correlation between greenspace usage at the data zone level and the probability for a data zone to record at least one Lyme disease case, either for Strava cycling trip per ha (OR = 0.73, 95% CI: 0.38-1.42, p = 0.36, Figure 3i, Table 1) or per capita (OR = 0.85, 95% CI: 0.48-1.49, p = 0.56, Figure 3j, Table 1).

Prediction 7: Greenspace usage index was negatively correlated with Lyme disease hazard within the 32 greenspaces (IRR = 0.23, 95% CI: 0.07-0.69, p = 0.01, Figure 3k, Table 1).

Prediction 8: There was no correlation between deprivation rank within 1 km of the 32 greenspaces and Lyme disease hazard (IRR = 0.75, 95% CI: 0.43-1.33, p = 0.33, Figure 3l, Table 1).

## Discussion

In this study, we found a strong, positive association between socioeconomic status and Lyme disease incidence in the city of Glasgow (UK), in line with previous research in North America,^5–7^ England and Wales.^8^ While several mechanisms could explain this effect (online supplementary figure 1), we tested two hypotheses relating to local landscape characteristics pre-exposure behaviour.

Our “landscape hypothesis” predicted that more advantaged areas have more high-risk habitat (woodlands);^7,21,22^ and that woodland cover is associated with Lyme disease incidence and environmental hazard. In full support of this hypothesis, more advantaged areas did have higher woodland cover at both the data zone spatial scale, and within a 1 km buffer around the 32 greenspaces surveyed. Woodland cover within data zones was positively associated with Lyme disease incidence, and woodland cover within the surveyed 32 greenspaces was positively associated with Lyme disease hazard. Woodlands are well documented to be high-risk habitats for Lyme disease,^14,15^ providing resources for key tick and pathogen hosts such as deer, rodents and birds, as well as microclimates suitable for tick survival and activity ^9,12,13^.

We found no direct correlation between deprivation rank within 1 km of greenspaces and Lyme disease hazard. This suggests that the environmental characteristics influencing infected tick density (tick and pathogen hosts, tick activity and survival), were broadly similar across our socioeconomic gradient. While we did not collect data on greenspace management, this finding does not support the idea of major differences in management in the surveyed areas, in ways that measurably impact ticks or pathogen. This is important as it demonstrates that more advantaged areas do not have higher risk per unit area of woodland, they simply have more woodland.

Our “behaviour hypothesis” predicted that people living in more advantaged urban areas are more exposed to Lyme disease pathogens due to higher usage of local greenspaces.^17^ We found no association between deprivation rank and our proxies of greenspace usage per unit area or per capita, either at the data zone level or across our 32 greenspaces. However, there might be deprivation-related disparities in the logging of cycling trips on Strava, or use of other social media platforms. Cycling data might also not completely reflect pedestrian usage of urban greenspaces. It remains possible that the patterns or types of use could differ,^16^ especially if there are concerns about safety, accessibility or poor-quality amenities in local greenspaces in deprived areas.^17–19^ This highlights the pressing need for robust urban footfall datasets that are available for researchers to use, which are currently lacking.

There could also be a deprivation-related disparity in non-local outdoor recreation. We had no information on where people acquired infection, only their residential postcode, so we do not know the proportion of Lyme disease cases contracted within or outside local greenspaces. While our landscape hypothesis was fully supported, suggesting a role of local woodland in explaining why Lyme disease incidence is higher in more advantaged areas, an unknown proportion of Lyme disease infections will have been acquired non-locally. In addition, people of higher socioeconomic status might spend more time outdoors in higher risk rural areas, such as national parks.^34^ This would likely substantially increase their exposure to infected ticks, because rural woodlands in the UK have higher environmental hazard of Lyme disease than wooded urban greenspaces.^29^

Interestingly, greenspaces with higher usage per unit area had lower environmental hazard of Lyme disease. Likely mechanisms explaining this include human (and dog [*Canis lupus familiaris*]) disturbance of wildlife that carry ticks,^35–39^ and more intensive management of these heavily used greenspaces such as frequent grass mowing and leaf clearing, which creates poor habitats for Lyme disease pathogen hosts such as rodents, and associated poor microclimate inhibiting tick survival and activity.^9,12,13^ This illustrates a key public health trade-off between environmental hazard and exposure: while the busiest urban greenspaces have the fewest infected ticks, thereby posing very low risk per person, the possibility of population-level risk should not be ignored.

To conclude, this study provides, to our knowledge, the first demonstration of socioeconomic disparities in Lyme disease while concurrently evaluating environmental and behavioural mechanistic pathways. The findings reveal a clear environmental justice issue: more advantage communities are disproportionately exposed locally to infected ticks because of higher local woodland cover (but not higher environmental hazard per unit area of woodland); meanwhile, more deprived areas have less woodland cover, so lower risk of Lyme disease but, simultaneously, this may mean fewer physical and mental health benefits associated with wooded greenspace. While we found no evidence that greenspace usage intensity (per unit area or per capita) is a mechanistic pathway linking socioeconomics with Lyme disease incidence, there remains the possibility of other behavioural pathways, such as types of greenspace usage, outdoor recreation in non-local higher risk rural environments, adoption of protective mitigation behaviours or medical seeking behaviours. Addressing these questions will be essential to fully understand the social drivers of Lyme disease and to develop effective public health responses.

## Data sharing

This study used aggregated anonymised Lyme disease data at data-zone level and did not involve identifiable individual participant data or direct involvement of human subjects. The disease case data at the data zone level will not be made available. All other datasets and R code used in this study will be made available after peer-review.

## Authors contributions

DJ, LG and RB initiated the project and secured funding; JH, LG and SLG designed the field methodology; SLG, JH and LG collected the data; GP conducted molecular analyses; SM and GP curated the Lyme disease case dataset; SLG analysed the data and generated figures; SLG and LG led the writing of the manuscript; All authors contributed to reviewing and editing the manuscript. All authors read and approved the final manuscript.

## Acknowledgements

We thank Noee Sellar and Patrycja Knapczyk for help with fieldwork. We also thank Public Health Scotland and NHS Highlands for granting us access to the Lyme disease case data. We would also like to thank Glasgow City Council for granting us access to sites used in our survey.

