## Supplementary materials for "When advantage turns into risk: disentangling landscape and behavioural drivers of socioeconomic inequality in Lyme disease risk, Glasgow as a case study"

**When advantage turns into risk: a multi-scale analysis to identify explanatory mechanisms of Lyme Disease**

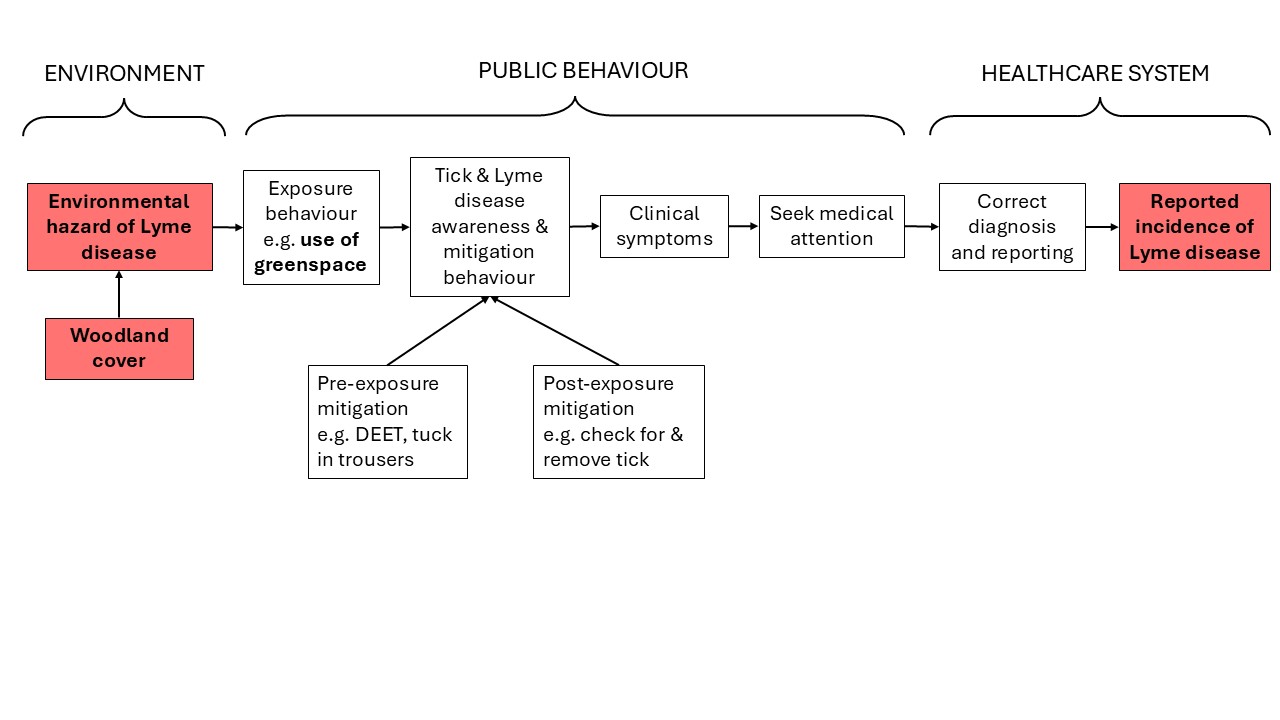

### Figure 1. Conceptual framework illustrating people’s exposure to tick-borne diseases.

This framework show environmental risk for tick-borne diseases, how people’s behaviour may impact their diagnosis (exposure risk, use of mitigation strategies, knowledge of clinical symptoms, seeking medical attention) and how the healthcare system leads to diagnosis and reporting. The boxes highlighted in red were investigated in the present study.

### Table 1: Indicators used to calculate the deprivation range and their description

| **Indicator** | **Description** |
| --- | --- |
| Income | Percentage and number of people who are income deprived |
| Employment | Percentage and number of people who are employment deprived |
| Health | Hospital stays related to alcohol or drug use  Standardised mortality ratio  Proportion of population being prescribed drugs for anxiety, depression or psychosis  Proportion of live singleton births of low birth weight  Emergency stays in hospital: standardised ratio |
| Education, skills and training | School pupil attendance  Attainment of school leavers  Working age people with no qualifications (standardised ratio)  Proportion of people aged 16-19 not participating in education, employment or training  Proportion of 17-21 years old entering university |
| Geographical access to services | Average drive time to a petrol station in minutes  Average drive time to a GP surgery in minutes  Average drive time to a post office in minutes  Average drive time to a primary school in minutes  Average drive time to a retail centre in minutes  Average drive time to a secondary school in minutes  Public transport travel time to a GP surgery in minutes  Public transport travel time to a post office in minutes  Public transport travel time to a retail centre in minutes  Percentage of premises without access to superfast broadband |
| Crime | Number of recorded crimes of violence, sexual offences, domestic housebreaking, vandalism, drugs offences, and common assault  Recorded crimes of violence, sexual offences, domestic housebreaking, vandalism, drugs offences, and common assault per 10,000 people |
| Housing | Number of people in households that are overcrowded  Number of people in households without central heating  Percentage of people in households that are overcrowded  Percentage of people in households without central heating |

### Supplementary text 1: Protocol used to test samples for *Borrelia burgdorferi* s.l.

DNA was extracted for each individual questing *I. ricinus* nymph by boiling each nymphs at 100 °C in 150 µl of 0.7 M NH_4_OH for 15 min. Tubes were then briefly centrifuged and heated at 100 °C for another 15 min with their lids open, until 70–100 µl of solution remained. The DNA was then used as a template in a real-time PCR targeting the 23S rDNA gene for *B. burgdorferi* s.l. The qPCR was implemented using Meridian Bioscience MyTaq Mix (Scientific Laboratory Supplies) in a CFX Connect real-time PCR detection system (Bio-Rad). Each reaction of 25 μl contained 12.5 μl of master mix, 1 μl of each primer at 10 pmol μl^−1^ (forward, 5′-CGAGTCTTAAAAGGGCGATTTAGT; reverse, 5′-GCTTCAGCCTGGCCATAAATAG), 1 μl of probe at 3.2 pmol μl^−1^ (5′-FAM-AGATGTGGTAGACCCGAAGCCGAGTG-TAMRA), 7.5 μl of nuclease free water and 2 μl of DNA extract using a program consisting of 5 min at 95 °C, followed by 40 cycles of 15 s at 95 °C and 1 min at 57 °C. A threshold cycle or *c*(*t*) value of 36 or lower was considered a positive result. Due to unforeseen circumstances, the original assay was switched for a protocol targeting ospA part way through the study. In this protocol, each reaction of 20 μl contained 10 μl of master mix, 1 μl of primers (forward, 5′-AATATTTATTGGGAATAGGTCTAA; reverse, 5′-CTTTGTCTTTTTCTTTRCTTACAA G)/probe (5′-FAM-AAGCAAAATGTTAGCAGCCTTGA-BHQ-1) mix, 6 μl of nuclease free water and 3 μl of DNA extract using a program consisting of 5 min at 95 °C, followed by 60 cycles of 5 s at 94 °C and 35 s at 60 °C, with a final incubation at 37 °C for 20 s. All other parameters and equipment remained unchanged. For every five DNA samples tested we included a negative (PCR grade water) control. A *c*(*t*) value of 40 or lower was considered a positive result. For both assays, each positive sample was viewed separately to ensure the quality and smoothness of the curve. A subset of previously tested samples (*n* = 282) was retested using the ospA assay to ensure result consistency, and the two methods produced identical results.

### **Table 2: Statistical model outputs**.

Full outputs from the GLMS and GLMMs including the prediction, level of analysis, response variable, covariate included, estimate, standard error, z-value and associated p-value for each covariate.

| Level | Response | Covariate | Estimate | Std. error | z-value | p-value |
| --- | --- | --- | --- | --- | --- | --- |
| **1-Effects deprivation on Lyme disease incidence** | | | | | | |
| Data Zone | Number of cases | Intercept | -11.48 | 0.34 | -33.73 | <0.001 |
|  |  | Deprivation rank | 0.21 | 0.09 | 2.25 | 0.02 |
|  |  | Year (Baseline: 2018) |  |  |  |  |
|  |  | 2019 | 0.24 | 0.41 | 0.59 | 0.55 |
|  |  | 2020 | 0.65 | 0.38 | 1.70 | 0.09 |
|  |  | 2021 | 0.97 | 0.36 | 2.69 | 0.007 |
|  |  | 2022 | 0.90 | 0.37 | 2.43 | 0.01 |
|  |  | 2023 | 0.69 | 0.38 | 1.83 | 0.07 |
|  |  | 2024 | 0.82 | 0.37 | 2.22 | 0.03 |
|  |  | Moran vector1 | -8.31 | 2.46 | -3.39 | 0.007 |
|  |  | Moran vector2 | 6.88 | 2.77 | 2.49 | 0.01 |
|  |  | Moran vector3 | -2.93 | 2.54 | -1.16 | 0.25 |
|  |  | Moran vector4 | 5.06 | 2.57 | 1.97 | 0.05 |
| **2-Effects of deprivation on woodland cover** | | | | | | |
| Data Zone | Deprivation index | Intercept | 7.62 | 0.32 | 240.58 | <0.001 |
|  |  | Woodland cover | -0.01 | 0.007 | -2.05 | 0.04 |
|  |  | Woodland cover² | 0.0005 | 0.0001 | 3.43 | <0.001 |
|  |  | Moran vector1 | -4.79 | 0.72 | -6.62 | <0.001 |
|  |  | Moran vector2 | 2.61 | 0.69 | 3.76 | <0.001 |
|  |  | Moran vector3 | 4.23 | 0.77 | 5.46 | <0.001 |
|  |  | Moran vector4 | -2.15 | 0.68 | -3.15 | 0.001 |
|  |  | Moran vector5 | -3.41 | 0.79 | -4.33 | <0.001 |
|  |  | Moran vector6 | -2.88 | 0.71 | -4.05 | <0.001 |
|  |  | Moran vector7 | 4.10 | 0.72 | 5.69 | <0.001 |
|  |  | Moran vector8 | -2.92 | 0.80 | -3.67 | 0.002 |
|  |  | Moran vector9 | 1.63 | 0.72 | 2.27 | 0.02 |
|  |  | Moran vector10 | 3.56 | 0.75 | 4.75 | <0.001 |
| 32 greenspaces | Deprivation index 1 km buffer | Intercept | 7.97 | 0.13 | 61.49 | <0.001 |
|  |  | Woodland cover 1 km buffer | 0.01 | 0.01 | 2.13 | 0.03 |
| **3-Effect of woodland cover on tick density/Lyme disease hazard** | | | | | | |
| 32 greenspaces | Number of infected nymphs | Intercept | -5.87 | 0.29 | -20.09 | <0.001 |
|  |  | Woodland cover | 0.78 | 0.37 | 2.09 | 0.04 |
|  |  | Temperature | -0.90 | 0.35 | -2.54 | 0.01 |
| **4-Effects of woodland cover on Lyme disease incidence** | | | | | | |
| Data Zone | Number of cases | Intercept | -8.32 | 0.09 | -89.86 | <0.001 |
|  |  | Woodland cover | -0.47 | 0.19 | -2.48 | 0.01 |
|  |  | Woodland cover² | 0.44 | 0.15 | 3 | 0.003 |
|  |  | Moran vector1 | -9.32 | 2.16 | -4.32 | <0.001 |
| **5-Are urban greenspaces used more in privileged areas?** | | | | | | |
| Data Zone | Strava trips per ha of greenspace | Intercept | 4.35 | 0.34 | 12.90 | <0.001 |
|  |  | Deprivation rank | -0.24 | 0.47 | -0.51 | 0.61 |
|  |  | Moran vector1 | 4.60 | 4.01 | 1.15 | 0.25 |
| Data Zone | Strava trips per capita | Intercept | -1.21 | 0.26 | -4.58 | <0.001 |
|  |  | Deprivation rank | -0.24 | 0.31 | -0.73 | 0.45 |
|  |  | Moran vector1 | 5.71 | 3.49 | 1.64 | 0.10 |
|  |  | Moran vector2 | 3.77 | 3.26 | 1.15 | 0.25 |
|  |  | Moran vector3 | -14.13 | 4.01 | -3.53 | <0.001 |
|  |  | Moran vector4 | -13.83 | 3.61 | -3.83 | <0.001 |
| 32 greenspaces | Parks’ usage | Intercept | 6.03 | 0.30 | 19.50 | <0.001 |
|  |  | Deprivation 1 km buffer | -0.50 | 0.37 | -1.34 | 0.2 |
| **6-The effects of urban greenspace usage on Lyme disease incidence** | | | | | | |
| Data Zone | Number of cases (binary) | Intercept | -8.44 | 0.21 | -40.90 | <0.001 |
|  |  | Strava trips per ha of greenspaces | -0.31 | 0.34 | -0.92 | 0.36 |
| Data Zone | Number of cases (binary) | Intercept | -8.42 | 0.20 | -41.9 | <0.001 |
|  |  | Strava trips per capita | -0.17 | 0.29 | -0.58 | 0.56 |
| **7-The effect of greenspaces’ usage on tick density/Lyme disease hazard** | | | | | | |
| 32 greenspaces | Number of infected nymphs | Intercept | -6.11 | 0.33 | -18.73 | <0.001 |
|  |  | Parks’ usage | -1.48 | 0.57 | -2.60 | 0.009 |
|  |  | Temperature | -0.99 | 0.35 | -0.84 | 0.005 |
| **8-The effects of deprivation on tick density/Lyme disease hazard** | | | | | | |
| 32 greenspaces | Number of infected nymphs | Intercept | -5.74 | 0.30 | -18.86 | <0.001 |
|  |  | Temperature | -1.07 | 0.35 | -3.02 | 0.002 |
|  |  | Deprivation 1 km buffer | -0.29 | 0.29 | -0.98 | 0.33 |
